# Clinical Evaluation of a COVID-19 Antibody Lateral Flow Assay using Point of Care Samples

**DOI:** 10.1101/2020.12.02.20242750

**Authors:** Won Lee, Steven Straube, Ryan Sincic, Jeanne A. Noble, Juan Carlos Montoy, Aaron E. Kornblith, Arun Prakash, Ralph Wang, Roland J. Bainton, Philip A. Kurien

## Abstract

**Introduction:** The ongoing SARS-CoV-2 pandemic has spurred the development of numerous point of care (PoC) immunoassays. Assessments of performance of available kits are necessary to determine their clinical utility. Previous studies have mostly performed these assessments in a laboratory setting, which raises concerns of translating findings for PoC use. The aim of this study was to assess the performance of a lateral flow immunoassay for the detection of SARS-CoV-2 antibodies using samples collected at PoC.

**Method:** One lateral flow immunoassay (Humasis^®^ COVID-19 IgG/IgM) was tested. In total, 50 PCR RT-PCR positive and 52 RT-PCR negative samples were collected at PoC. Fifty serum specimens from Dec 2018 to Feb 2019 were used as controls for specificity. Serum samples collected between Dec 2019 to Feb 2020 were used as additional comparators. Clinical data including symptom onset date was collected from patient history and the medical record.

**Results:** The overall sensitivity for the kit was 74% (95% CI: 59.7% -85.4%). The sensitivity for IgM and IgG detection >14 days after date of onset was 88% (95% CI: 68.8% -97.5%) and 84% (95% CI: 63.9% – 95.5%), with a negative predictive value (NPV) of 94% for IgM (95% CI: 83.5% - 98.8%) and 93% for IgG (95% CI: 81.8% - 97.9%). The overall specificity was 94% (95% CI: 83.5% - 98.8%). The Immunoglobulin specific specificity was 94% for IgM (95% CI: 83.5% - 98.8%) and 98% for IgG (95% CI: 89.4% - 100.0%), with a positive predictive value (PPV) of 88% for IgM (95% CI: 68.8% - 97.5%) and 95% for IgG (95% CI: 77.2% - 99.9%) respectively for samples collected from patients >14 days after date of onset. Specimen collected during early phase of COVID-19 pandemic (Dec 2019 to Feb 2020) showed 11.8% antibody positivity, and 11.3% of PCR-negative patients demonstrated antibody positivity.

**Discussion:** Humasis^®^ COVID-19 IgG/IgM LFA demonstrates greater than 90% PPV and NPV for samples collected 14 days after the onset of symptoms using samples collected at PoC. While not practical for the diagnosis of acute infection, the use of the lateral flow assays with high specificity may have utility for determining seroprevalence or seroconversion in longitudinal studies.

## INTRODUCTION

The outbreak of coronavirus disease 2019 (COVID-19), caused by the severe acute respiratory syndrome coronavirus 2 (SARS-CoV-2), continues to place unprecedented strain on the healthcare system. By October 2020, more than 40 million confirmed cases and one million related deaths have been reported worldwide [1]. While governmental interventions to slow viral spread have been effective in certain parts of the world, resurgence in new cases and deaths in the United States demonstrate that strategies to reopen businesses and ease restrictions on human mobility and interactions will depend on accurate estimates of population level infection and markers of immunity [2].

The confirmatory diagnosis of COVID-19 infection largely depends on molecular techniques. The current gold standard is the detection of viral RNA in the respiratory tract through assays using real-time reverse transcriptase polymerase chain reaction (RT-PCR) [3]. The PCR test, however, has a limited time window for highest sensitivity, and may not adequately capture recent infection, or be informative of previous exposures [4]–[6]. Therefore, serologic assays, which detect serum antibodies to SARS-CoV-2, may be necessary to complement PCR-based techniques especially to characterize the population-based prevalence of infection and immunity [7].

There are two main ways to detect serum antibodies against SARS-CoV-2. Enzyme-linked immunosorbent assay (ELISA), a laboratory method for quantitative antibody detection, is an established method but has long turn-around time and a high cost burden. Immunochromatographic lateral flow assays (LFA), on the other hand, can be used as point of care (PoC) assays and typically produce results in minutes, albeit results that are qualitative and not quantitative.

Since the United states government established an Emergency Use Authorization for COVID-19 related PoC tests in February 2020, more than 100 manufacturers have marketed COVID-19 LFA antibody kits. However, questions and concerns remain about the validity and diagnostic performance of these PoC LFA antibody tests and previous studies have demonstrated a varying degree of tests’ sensitivity and specificity [8]–[11]. Furthermore, these studies have been performed in a laboratory setting, which may not translate for use as a PoC test in clinical practice.

The aim of this study was to critically evaluate the diagnostic performance of a COVID-19 PoC LFA antibody test (Humasis^®^ LFA) to assess its utility as a point of care assay.

## METHODS

### Ethical Approvals

This study was approved by the institutional review board at the University of California, San Francisco (UCSF) and Zuckerberg San Francisco General Hospital (ZSFG).

### Study Design

Specimens were collected from patients tested for Sars-CoV-2 and bio-banked controls. Sars-CoV-2-positive and negative samples were obtained at bedside from a convenience sample of patients who underwent SARS-CoV-2 real-time polymerase chain reaction (RT-PCR) testing of nasopharyngeal and/or oropharyngeal swabs in the emergency department (ED), perioperative arena, or inpatient wards. Both symptomatic and asymptomatic patients were included. Symptomatic patients were asked of their symptom onset date. One specimen per individual was included. A total of 52 SARS-CoV-2 PCR positive and 52 PCR negative patients were included. The majority (n = 40) of PCR negative samples were collected in the perioperative arena. These patients underwent PCR tests as a part of their routine preoperative assessment. At the time of sample collection, PCR negative patients were asked if they had any history of upper respiratory tract infection (URI) symptoms or other symptoms concerning for COVID-19 since February of 2020.

To assess specificity, we included 50 bio-banked plasma specimens collected between Dec 2018 and Feb 2019 that were presumed to be COVID-19 negative due to the interval between collection and the onset of the pandemic. We also tested 51 serum specimens collected between Nov 2019 through Jan 2020. Testing of bio-banked samples was performed in the laboratory setting.

Patients with immune deficiency (innate or acquired) were excluded from enrolling in our study as these patients may not generate the necessary immune response to the viral infection. Data obtained from two specimens that did not conform to our study design were excluded. These samples were incubated for longer than the time allotted in the protocol prior to reading the results. Clinical data including medical history, patient-reported symptom onset date, and PCR results were collected from electronic health records and stored in a HIPAA-secure database.

### Sample Collection and Preparation

Patient blood samples in the clinical arena were collected and applied to the kits at the patients’ bedside. These samples were collected at PoC by either using existing intravenous (IV) lines or by finger lancet. If samples were to be collected from an existing IV-line, study providers were instructed to draw an appropriate amount of volume of blood and discard it prior to collecting samples to minimize sample dilution. Blood samples from the finger lancet could be applied directly to the sample well or pipetted through a manufacturer provided micro-pipette, prior to transferring to the sample well.

Pre-COVID19 plasma samples were thawed at room temperature before using a micro-pipette to apply the sample to the lateral flow assay cartridge using sterile technique. All sample handling followed UCSF biosafety committee-approved practices.

### Immunochromatographic Lateral Flow Assay (LFAs)

Lateral flow assays were used per manufacturer instructions. At least 10 microliters of blood or plasma sample was transferred to the indicated sample well, followed by the manufacturer provided diluent. The lateral flow cartridges were incubated for 15 minutes at room temperature before reading. Each cartridge was assigned a unique number by the study member responsible for sample collection. For analysis, two clinician assessors blinded to the specimen’s PCR status read the result on a cartridge and assigned a binary score (0 for negative, 1 for positive) for immunoglobulin M (IgM) and immunoglobulin G (IgG) antibodies; a third adjudicated disagreements by assigning a binary score. Per the manufacturer guidelines, test line intensity did not factor into our analysis; any visible band was considered positive. All the cartridges used in testing were from the same lot.

### Statistical Analysis

Cases were divided by time interval; (1) all time points (2) 7 days and beyond from onset (3) 14 days and beyond from onset. Onset is defined as a date from symptom onset or PCR date for asymptomatic patients. Sensitivity was based on results in PCR positive samples. Specificity was based on results in pre-SARS-CoV-2 samples. Stratified analysis was performed on symptomatic patients using the same time interval. Binomial exact 95% confidence intervals were calculated for all estimates. Two assessors’ scores were used to calculate Cohen’s Kappa for inter-reader agreement statistics. Data was aggregated and processed in Python (Pycharm®, Prague, Czech Republic), and subsequent analyses were conducted using STATA 15.1 (College Station, TX).

## RESULTS

### Population Demographics

The 50 SARS-CoV-2 PCR-positive patients ranged from 21 to 93 years of age with median age of 47.5 years (interquartile range [IQR]: 35 – 62). SARS-CoV-2 PCR-negative patients ranged from 18 to 90 years of age with median age of 53 years (IQR: 35 – 70). Forty percent of PCR positive and 46% of PCR negative patients were female. Seventy-six percent of PCR-positive and 33% of PCR-negative patients reported symptoms. 96% of the PCR-positive patients were hospitalized and 6% were admitted to the ICU. The median of days from onset was 14 days at time of sample collection for PCR positive patients (IQR: 7 – 24 days) (Table 1).

**Table 1:**
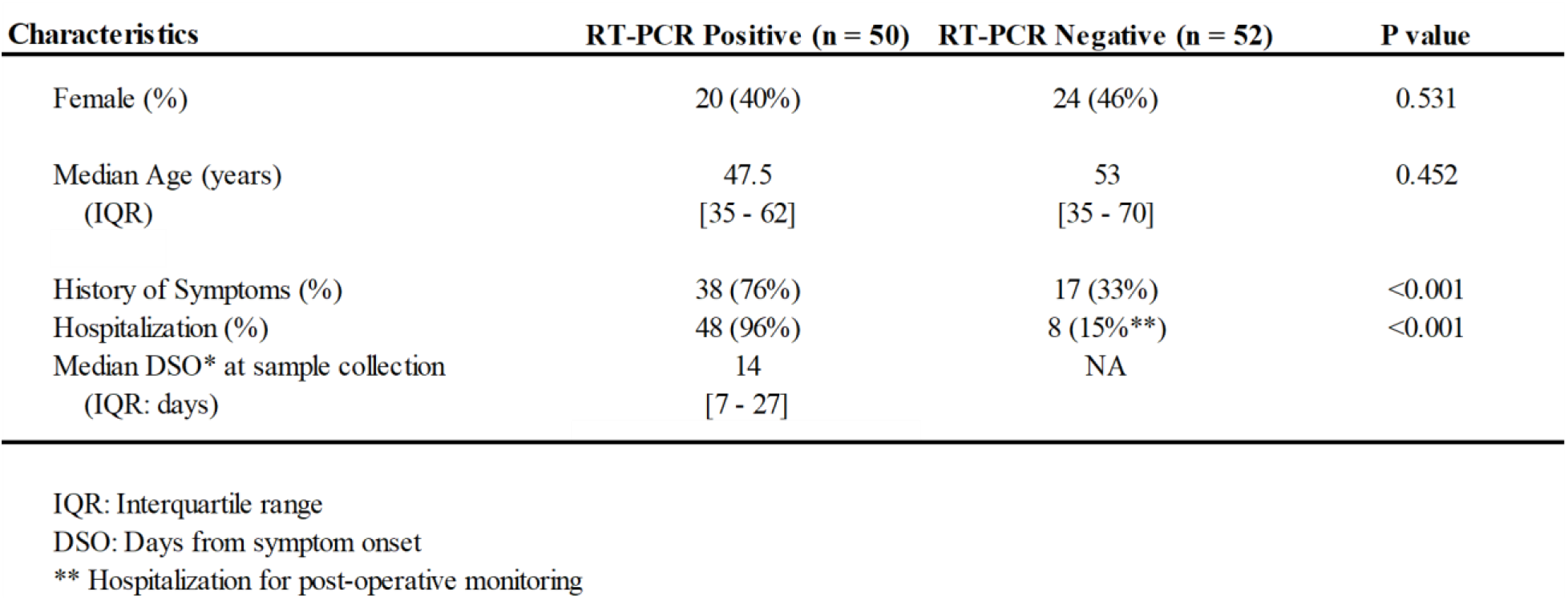
Baseline demographic characteristics, median age, presence of symptoms, proportion of patients who required hospitalization, and proportion of symptomatic patients with their median days from symptom onset at sample collection for serologic testing. Only one sample per patient was included in the study.

### Performance Characteristics

Sensitivities and specificities obtained are summarized in **Table 2**. The overall sensitivity for the kit was 74.0% (95% confidence interval [CI]: 59.7% - 85.4%) and rose with increasing time from the date of symptom onset. Samples taken from patients with a symptom onset of less than 7 days showed 50.0% sensitivity (n = 6; 95% CI: 11.8% - 88.2%), while those beyond 7 days rose to 81.5% (n = 38; 95% CI: 65.7% - 92.3%) and samples from 14 days and beyond demonstrated 88.0% (n = 25; 95% CI: 68.8% - 97.5%) sensitivity. The analysis on symptomatic patients showed increased sensitivity; sensitivities of symptomatic patient samples of greater than 7 and 14 days from the symptom onset date were 82.9% (n = 35; 95% CI: 66.4% - 93.4%) and 91.3% (n = 23; 95% 72.0% - 98.9%), respectively.

**Table 2a:** Overall summary statistics for immunochromatographic lateral flow assay (LFA).

Overall sensitivity and specificity of Humasis LFA compared to RT-PCR as a gold standard
| LFA Antibody result (Index Test) | RT-PCR<br>(Reference Standard) |  | Total |
| --- | --- | --- | --- |
|  | Positive | Negative |  |
| **Positive** | 37 | 3 | 40 |
| **Negative** | 13 | 47 | 60 |
| **Total** | 50 | 50 | 100 |

**Table 2b:** Samples are categorized by time from onset, defined as time (in days) from patient-reported symptom onset or RT-PCR date for asymptomatic patient, to sample collection date. Percent seropositivity is reported with 95% confidence intervals (95% CI). The IgM refers to immunoglobulin M, and IgG refers to immunoglobulin G. Specificity is determined relative to pre-COVID-19 negative control serum samples.

|  | **Days from RT-PCR/Symptom Onset (Sensitivity %; 95% CI)** |  | **All Patients % (95% CI)**<br>**(n = 50)** |
| --- | --- | --- | --- |
|  | Greater than 7 days | Greater than 14 days |  |
| **All patients** | (n = 38) | (n = 25) |  |
| IgG or IgM | 81.5% (65.7% - 92.3%) | 88.0% (68.8% - 97.5%) | SE: 74.0% (59.7% - 85.4%)<br>SP: 94.0% (83.5% - 98.8%) |
| IgG | 76.3% (59.8% - 89.6%) | 84.0% (63.9% - 95.5%) | SE: 70.0% (55.4% - 82.1%)<br>SP: 98.0% (89.4% - 100%) |
| IgM | 81.5% (65.7% - 92.3%) | 88.0% (68.8% - 97.5%) | SE: 74.0% (59.7% - 85.4%)<br>SP: 94.0% (83.5% - 98.8%) |
| **Symptomatic Patients** | (n = 35) | (n = 23) |  |
| IgG or IgM | 82.9% (66.4% - 93.4%) | 91.3% (72.0% - 98.9%) | SE: 81.6% (65.7% - 92.3%)<br>SP: 94.0% (83.5% - 98.8%) |
| IgG | 77.1% (59.9% - 89.6%) | 87.0% (66.4% - 97.2%) | SE: 76.3% (59.8% - 88.6%)<br>SP: 98.0% (89.4% - 100%) |
| IgM | 82.9% (66.4% - 93.4%) | 91.3% (72.0% - 98.9%) | SE: 81.6% (65.7% - 92.3%)<br>SP: 94.0% (83.5% - 98.8%) |

SE: Sensitivity SP: Specificity CI: Confidence Interval

The true positive rate did not change by combining IgM and IgG results; sensitivity by IgM alone mirrored overall sensitivity. The sensitivity by IgG was less than that for IgM+IgG; 76.3% (95% CI: 59.8% - 89.6%) and 84.0% (95% CI: 63.9% - 95.5%) for samples beyond 7 and 14 days, respectively. Including only samples from symptomatic patients increased the IgG sensitivity to 77.1% (beyond 7 days; 95% CI: 59.9% - 89.6%) and 87.0% (beyond 14 days; 95% CI: 66.4% - 97.2%). The negative predictive value (NPV) for IgM was 94% (95% CI: 83.5% - 98.8%) and for IgG was 93% (95% CI: 81.8% - 97.9%).

The overall test specificity in pre-COVID-19 samples was 94.0% (95% CI: 83.5% - 98.8%), with 94.0% (95% CI: 83.5% - 98.8%) and 98.0% (95% CI: 89.4% - 100.0%) for IgM and IgG specific specificity. The positive predictive value (PPV) was 88% (IgM; 95% CI: 68.8% - 97.5%) and 93% (IgG; 95% CI: 89.4% - 100.0%) for samples collected from patients with >14 days after symptom onset.

Kappa agreement between assessors was 0.94 for IgM and 0.98 for IgG. There were no invalid tests that required repetitive sampling.

### Time Point Comparison

We sampled 50 pre-COVID bio-banked controls (from December 2018 to February 2019) and another 51 early-COVID era bio-banked samples (from December 2019 to February 2020), when the incidence of infection locally may have been on the rise. Samples from PCR negative patients were collected between May and August of 2020.

For early-COVID bio-banked samples, we identified 11.8% (95% CI: 4.5% - 23.9%) of samples being either IgG/IgM positive, with IgM and IgG antibody positivity of 11.8% and 4.0%, respectively. From 52 SARS-CoV-2 PCR-negative samples, we identified 11.3% (95% CI: 4.2% - 23.0%) of samples being either IgG/IgM positive. There was a non-statistically significant difference for patients who reported having URI or other symptoms concerning for COVID-19 since the outbreak. Among PCR-negative patients, 17.6% of samples from patients with a history of symptoms demonstrated antibody positivity (95% CI: 5.3% - 45.2%) compared to just 5.7% (95% CI: 1.4% - 21.0%) from patients who did not report previous symptoms (Figure 1).

**Figure 1:**
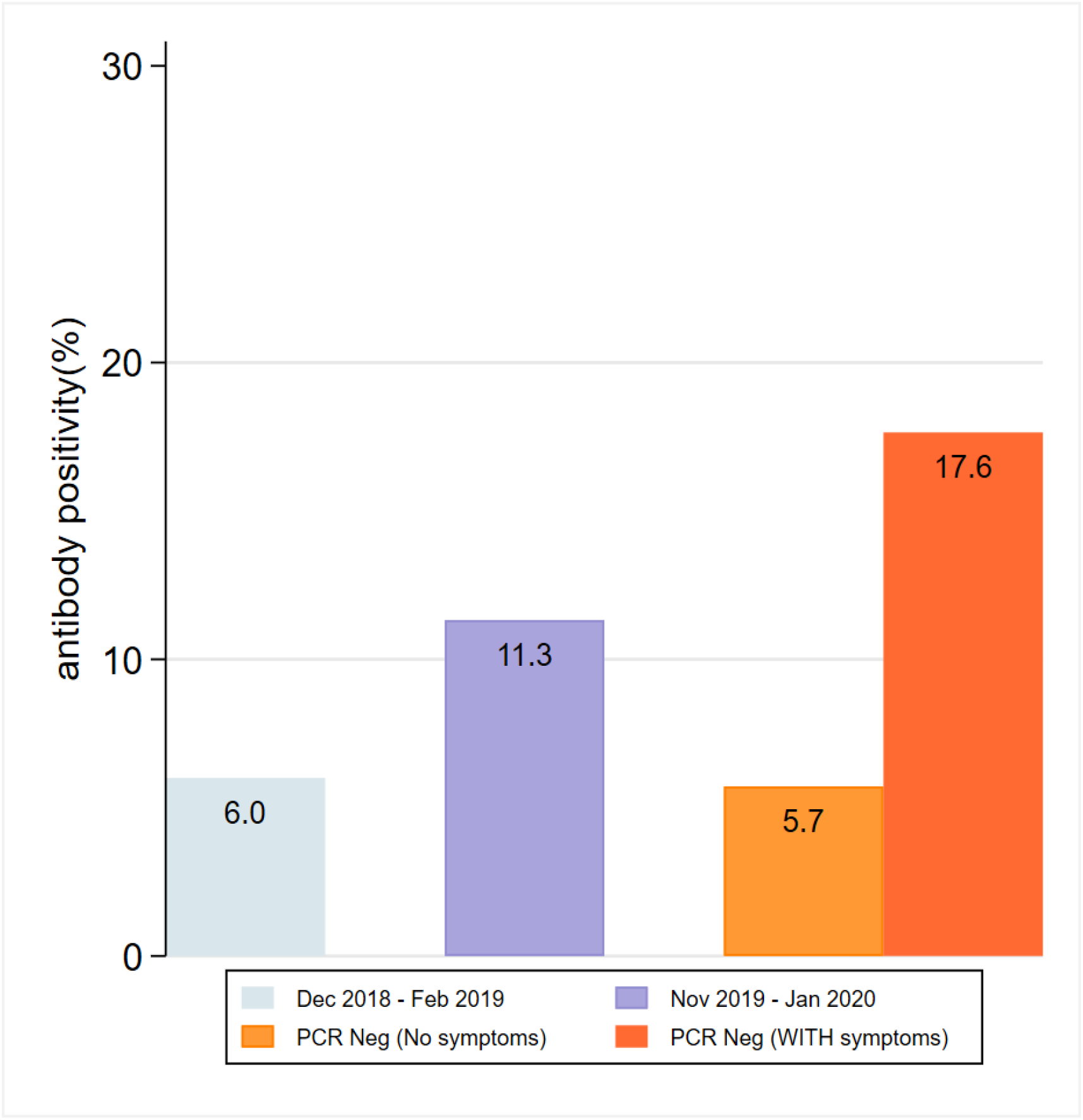
Antibody positivity noted on samples from November 2019 to January 2020 (bio-banked samples, unknown symptom history) compared to samples collected at PoC from PCR negative patients. PCR negative patients are divided by symptom history. Dec 2018 - Feb 2019 bio-banked samples, which were used for specificity test of the study. Results are not statistically significant.

## DISCUSSION

We evaluated the performance of a LFA assay that has not been previously tested in the acute-care setting in the US. As many devices are released to the market, it is imperative that each undergoes a thorough vetting to ensure adequate test performance for its intended clinical use. We found good clinical specificity (94%) using pre-COVID samples. Clinical sensitivity from 53 whole blood COVID-19 positive samples demonstrated improvement as the time interval from symptom onset to diagnostic test increased, with the highest sensitivity from samples collected 14 days from symptom onset or after. This is concordant with previous studies evaluating the performance of other LFA devices [8]–[11].

Seroconversion for IgM and IgG for COVID-19 infections occurs on average 2 – 3 weeks after the initial infection, and it can occur simultaneously or sequentially [12]. Levels of IgM may begin to wane as early as the 3^rd^ week after infection, and for this reason LFAs that can simultaneously measure IgM and IgG are considered more useful in detecting seroconversion [13]. Whitman et al. found combining IgM and IgG results modestly increased sensitivity values in their study [8]. Although we did not find that the combination improved sensitivity, the median time of testing in our study was 14 days after symptom onset. Increased sampling of patients with a longer interval after symptom onset or infection date might have altered this finding.

We also did not take band strength into consideration. LFAs are meant as qualitative tests and the manufacturer insert specifies that the presence of a band at any strength should be considered as a positive result. Nevertheless, the noticeable variance in band strength does raise a question of whether it correlates with the serum concentration of protective antibodies. Ibarrondo et al. propose that there is a rapid decay of anti-SARS-CoV-2 antibodies, especially for patients with mild symptoms [14] thus suggesting that quantification of the band may have implications for residual immunity. While our study was not powered to compare the level of symptoms, future studies should consider correlating band strength on LFAs with symptom strength or illness severity. A previous study has shown that a suboptimal antibody response may actually promote pathology, resulting from an antibody-dependent enhancement phenomenon [15]. Again, correlating quantitative serum antibody level to possibility of reinfection may be an important avenue for future studies.

Given the time trajectory of antibody formation after onset of infection, these serological tools should not, and cannot replace PCR-based testing for diagnosis of acute infection. Both the Centers for Disease Control and the Public Health Agency of Canada maintain that serologic assays should not be used as an aid in the diagnosis of acute infection [16], [17]. As such, we do not think that these devices have a role in the clinical patient setting. There are other methods of antibody testing available with higher fidelity, including serum IgG ELISAs, albeit at higher cost [18]. These tools, however, could be useful for determining community level immunity, either due to infection itself or after vaccination, especially in areas where access to more sophisticated and expensive quantitative analysis may be limited.

Finally, the main concerns with these PoC LFA kits involves the subjective nature of running the assays, including quantity/volume of sample loaded and assessment of sample test lines. To our knowledge, this was the first evaluation study of these LFA devices at point of care using whole blood drawn at the patient’s bedside. Our study investigators noted difficulty with drawing the recommended 10 microliter of sample volume from the finger lancet, at times, requiring more than one attempt. Multiple draws meant that more than 10 microliters of sample may have been applied to the cartridge. Also, the subjective nature of assessing whether a band is present makes it difficult for standardized use among global healthcare professionals with good reproducibility. It may be prudent for manufacturers to provide an automatic tool that could replace human readers for diagnosis using these kits.

### Limitations

A few study limitations should be noted. Notably, our samples were collected from the San Francisco Bay Area. Different strains of SARS-CoV-2 exist around the world [19], and whether our result will be reproducible for other strains is unknown [20]. A second limitation is that the samples used to evaluate the specificity as true-negatives were from bio-banked plasma samples and were carried out in a controlled laboratory setting. Also, a majority of our PCR positive patients were admitted to the hospital, which limits generalizability. Lastly, we have compared the LFA test results to those of RT-PCR. While PCR test remains the molecular gold standard for COVID-19 diagnosis, PCR tests, themselves, are subject to inherent diagnostic uncertainties [21], and comparing the presence of viral RNA in oral or nasopharyngeal secretions to serum antibodies has drawbacks. Therefore, our findings may not truly reflect false positives and false negatives, which would be better further validated with other molecular antibody tests, such as serum IgG ELISAs. Ng et al. have identified SARS-CoV-2 S-reactive IgG antibodies from May 2019 samples, representing preexisting humoral immunity from exposure to other coronavirus strains [22]. ELISA assays may have helped to better differentiate if false positives were due to existence of the antibodies. Similarly, false negative data points may be observed due to the failure or a delay in mounting antibody responses. For purposes of this study, these variables would not likely change conclusions about the utility of LFAs in acute infectious phase testing.

## Conclusions

The Humasis® COVID-19 IgG/IgM LFA had a sensitivity and specificity of 74% and 94%, corresponding to a greater than 90% NPV for samples collected 14 days after the onset of symptoms. While not useful for diagnosis of acute infection, the use of the lateral flow assays in determining seroprevalence or seroconversion in longitudinal studies may be useful.

## Data Availability

Data available at request

## Acknowledgement

We thank Dr. Matthias Braehler and Sara Nedkov for their support of this project. We thank the manufacturer of Humasis kits for donation of test kits.

